# From waistlines to death rates: a 30-year conversation between obesity and hepatocellular carcinoma

**DOI:** 10.1101/2025.08.01.25332662

**Authors:** Jie Xiong, Cheng Liu, Amir Muhammad, XiangCheng Xiao, Mingmei Liao

**Affiliations:** Department of Nephrology, Xiangya Hospital, Central South University, Changsha, Hunan, China; Department of Nuclear Medicine, Xiangya Hospital, Central South University, Changsha, Hunan, China; National Health Commission (NHC) Key Laboratory of Nanobiological Technology, Xiangya Hospital, Central South University, Changsha, Hunan, China

## Abstract

**Background:** The global rise in obesity has led to an increasing burden of hepatocellular carcinoma (HCC) associated with high BMI, alongside viral hepatitis. This study evaluates the global burden of HCC due to high BMI, using the Global Burden of Disease (GBD) framework.

**Methods:** Data from GBD 2021 were analyzed to compare age-standardized death rates (ASDR) and disability-adjusted life-years (ASR) of DALYs for high BMI-associated HCC from 1990 to 2021. The Estimated Annual Percentage Change (EAPC) was calculated using linear regression, and the Nordpred model was applied for projections from 2022 to 2040.

**Results:** From 1990 to 2021, both ASDR and ASR of DALYs for high BMI-associated HCC showed significant increases. The burden was higher in males, older age groups (especially 80+), and regions with intermediate SDI values. Projections suggest that the global burden will continue rising until 2040 but at a slower rate, potentially stabilizing by then.

**Conclusion:** The global burden of high BMI-associated HCC is increasing, especially in males and the elderly. Tailored prevention and treatment strategies are crucial to address regional trends and mitigate the growing disease burden.

## 1. Introduction

Liver cancer, especially hepatocellular carcinoma (HCC), represents a major global health burden. According to epidemiological statistics, its incidence ranks as the sixth most prevalent malignant tumor worldwide, while it is the third highest cause of cancer-related mortality [1–2]. For decades, chronic infection with hepatitis B virus (HBV) and hepatitis C virus (HCV) have been recognized as the primary etiological drivers of hepatocarcinogenesis[3–4], maintaining central importance in the development of strategies and the allocation of healthcare resources for the prevention and treatment of liver cancer.

In recent years, the growing problem of obesity has been reshaping the public health landscape[5–6]. The Body Mass Index (BMI) has been widely used in public health surveillance as a standardized indicator to assess overweight and obesity. With the rapid increase in the proportion of overweight and obese people worldwide, more than one-third of the population is now affected, posing a grave threat to public health and a substantial economic burden[7]. Numerous studies have shown that BMI is significantly associated with the occurrence and progression of a variety of tumors, especially in patients with prostate cancer[8], breast cancer[9] and colorectal cancer[10–11]. High BMI is also consistently associated with higher cancer mortality, which not only exacerbates the burden on the public health system, but also poses new challenges for optimizing existing cancer prevention and control strategies. Meanwhile, Wu et al.[12] revealed a strong association between high BMI and hepatocellular carcinoma (HCC). The study pointed out that the risk of liver cancer in overweight individuals was 1.4 to 4.1 times higher than that of the general population. By 2021, the number of deaths from primary liver cancer due to high BMI had climbed to about 60,000 and continues to increase. This trend has become an emerging challenge for the global public health system and has had a severe impact on existing medical resources. Nevertheless, the lack of a systematic assessment of the global burden of HCC associated with high BMI has limited the development of precise, evidence-based health policies by the World Health Organization and governments to effectively address evolving medical needs in the future [13].

This study utilized the data on high BMI-associated hepatocellular carcinoma (HCC) from the GBD 2021 study to systematically evaluate temporal trends in the disease burden by sex, age group, and geographic region between 1990 and 2021, and further predicted future trends through the Nordpred model. The primary aim was to generate robust, theory-driven evidence to inform targeted interventions and support the precise formulation and effective implementation of global public health policies addressing high-BMI–associated HCC. Concurrently, by offering a detailed, evidence-based framework tailored to China, this work seeks to optimize national liver cancer prevention and control strategies, strengthen public health infrastructure, and ultimately mitigate the escalating burden of high BMI–associated HCC on healthcare resources.

## 2. Materials and methods

### 2.1 Data source

The data for this study were sourced from the Global Burden of Disease (GBD) 2021 database, which encompasses 204 countries and territories grouped into 21 regions and 7 super-regions. Its cancer framework encompasses all malignancies except for specific cancer types (e.g., nonmelanoma skin cancers, benign and in situ cancers, etc.) and select hematopoietic cancers. The database details cancer incidence, mortality, and risk-factor data disaggregated by sex, region, country, and exposure through the GBD Results Tool (https://ghdx.healthdata.org/gbd-results-tool). In addition, the GBD 2021 provides base statistics for males, females, and all genders for each of the 23 age groups from birth to 95 years and older[14–15]. Given the extremely low burden of high BMI–associated HCC in individuals under 20 years, our analysis focused on those aged ≥20 years, subdivided into 16 five-year age groups (20–24, 25–29, …, 90–94, and ≥95 years) [16–17]. We systematically extracted and calculated ASDR and ASR of DALYs for high BMI-associated HCC globally from 1990 to 2021. All rates were standardized to the 2021 global standard population age structure.

### 2.2 Sociodemographic Index (SDI)

The sociodemographic index (SDI) serves as a composite measure that reflects a country’s level of social and economic development[18]. This summary measure includes a nation’s economy as measured by lag-distributed income (LDI) per capita, mean education for those 15 and older, and the total under 25 fertility rates of nations. This index is used in the Global Burden of Disease studies because the outcomes measured by the SDI correlate strongly with health outcomes. Based on the country’s SDI score, each country has been classified into one of the five categories—high, high– middle, middle, low–middle, and low.

### 2.3 Statistical analysis

First, in order to gain a comprehensive understanding of the burden of disease in high BMI-associated HCC, this study conducted a multilevel analysis. Detailed comparisons of ASDR and ASR of DALYs from 1990 to 2021 were conducted at the global, regional (21 GBD regions), and national (204 countries and territories) levels. Additionally, comparisons were conducted across the five SDI quintiles (high, high-middle, middle, low-middle, and low). Descriptive analyses were performed to visualize the trends and characterize the differential characteristics of the high BMI-associated HCC burden.

Second, to more precisely quantify the evolutionary dynamics of the disease burden, this study employed a linear regression model to calculate the Estimated Annual Percentage Change (EAPC) value and conducted a hierarchical cluster analysis based on this value [19]. This approach enabled a deeper assessment of the disease burden change patterns in each GBD region, allowing the identification of regions with similar trends. The 21 GBD regions were classified into four categories, representing different scenarios— ranging from significant increase and slight increase, remaining stable or slight decrease to significant decrease.

Finally, this study utilized the Nordpred model to project the global disease burden of high BMI-associated HCC from 2022 to 2040. The Nordpred model combines the age-period-cohort (APC) model with disease burden prediction methodologies, enabling a detailed analysis of the effects of age, period, and cohort factors on the risk of disease incidence and mortality. This approach facilitates scientifically grounded projections of future disease burden [20–22].

In the process of data processing and analysis, the R software program (version 4.2.3) was used in this study to organize, analyze, and visualize the data. Differences were considered statistically significant when the P value was less than 0.05.

### 2.4 Ethical Considerations

All data for this study were derived from the GBD 2021 database. The University of Washington Institutional Review Board granted a waiver of informed consent for the use of these de-identified data. Therefore, no additional ethics committee approval was required for this study. This study follows the Guidelines for Accurate and Transparent Health Estimates Reporting (GATHER).

## 3. Results

### 3.1 Global trend of high BMI-associated hepatocellular carcinoma

Globally, the disease burden of high BMI-associated hepatocellular carcinoma has increased significantly, with marked geographical disparities. Regions with the highest burden were predominantly concentrated in North America (including parts of the Nordic region, the United States, and Canada), selected European countries (e.g., Finland, Germany), East and South Asia, and parts of Africa. In these areas, ASDR ranged from 1.13 to 9.54 per 100,000, and ASR of DALYs ranged from 31.94 to 243.77 per 100,000, both significantly higher than the global average (P<0.01). In contrast, regions with lower disease burdens included parts of Southeast Asia (e.g., Myanmar, Vietnam, Cambodia) and South American countries such as Argentina, with ASDR below 0.29 per 100,000 and ASR of DALYs under 7.23 per 100,000 (Fig 1).

**Fig 1.**
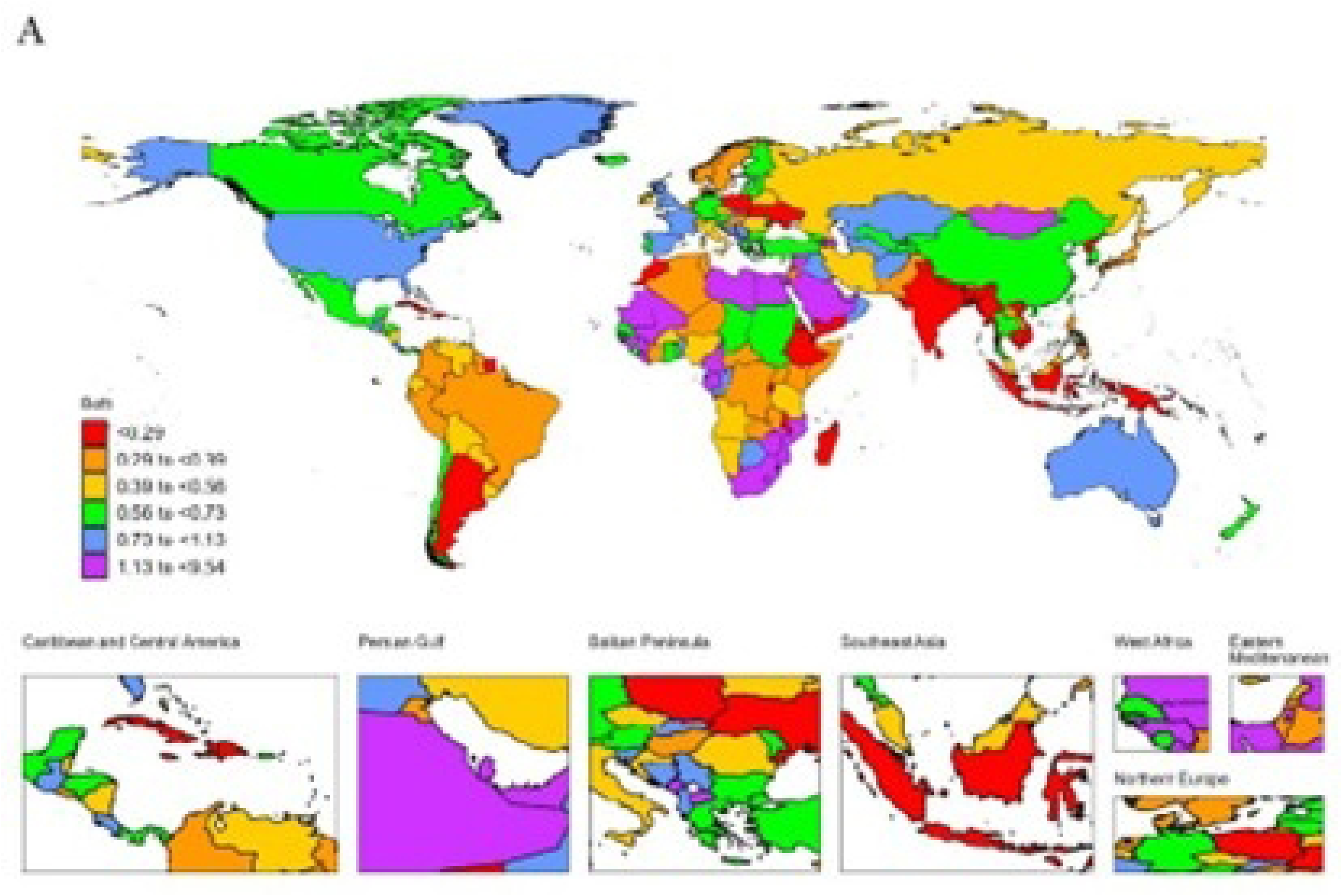
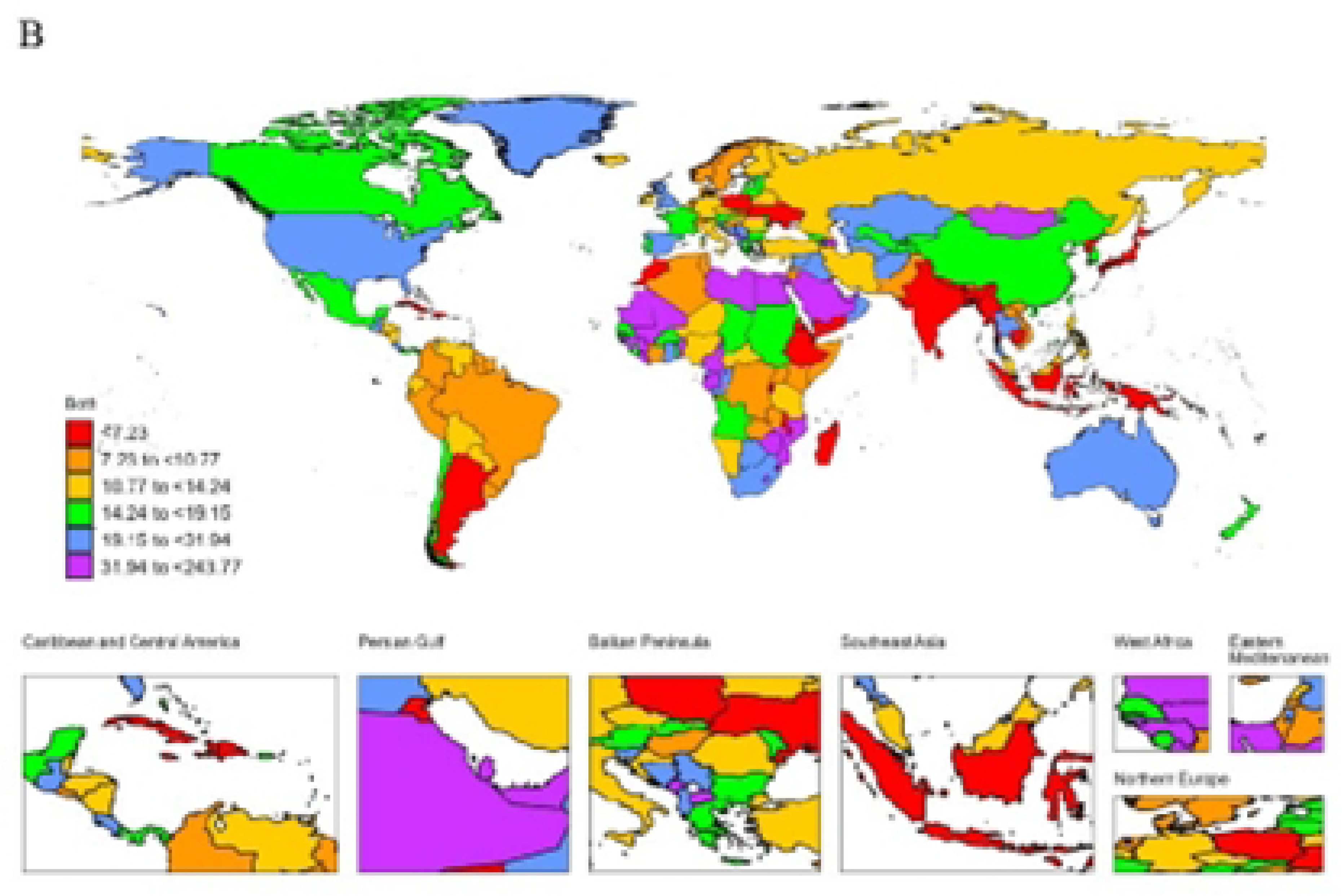
Global trends in ASDR (A) and ASR of DALYs (B) of high BMI-associated HCC. Abbreviations: ASDR,Age-standardized death rate; ASR,age-standardized rate; DALYs, disability-adjusted life-years.

### 3.2 Trends of high BMI-associated hepatocellular carcinoma by regions

At the regional level, the burden of high BMI-associated HCC has significantly increased in most regions. Notably, the rate of increase was most pronounced in the Oceania and South Latin America regions. In Oceania, the ASDR rose at an annualized rate of 4.81% (95% CI: 4.64-4.97), and the ASR of DALYs increased at a rate of 4.64% (95% CI: 4.50-4.78). Similarly, in South Latin America, the growth rates of ASDR and ASR of DALYs reached 4.21% (95% CI: 4.21-4.71) and 4.30% (95% CI: 4.04–4.56), respectively. Additionally, East Asia (ASDR of 3.87%, 95% CI: 3.73-4.01) and high-income North America (ASDR of 3.92%, 95% CI: 3.66-4.17) also exhibited substantial upward trends. In contrast, high-income Asia-Pacific was the only region to show a significant declines in both the ASDR (-0.39%, 95% CI: -0.82-0.05) and ASR of DALYs (-0.89%, 95% CI: -1.29 to -0.49). These findings suggest that the burden of high BMI-associated HCC is increasing most rapidly in resource-abundant regions (e.g., Oceania) and regions with emerging economies (e.g., East Asia, South Latin America), while a negative growth trend was observed in high-income Asia-Pacific regions (Table 1).

**Table 1.** Regional distribution of high BMI-associated HCC burden in both sexes for ASR, 1990–2021.

|  | location | measure | EAPC | LCI | UCI | EAPC(95%CI) |
| --- | --- | --- | --- | --- | --- | --- |
| 1 | East Asia | Deaths | 3.866077345 | 3.726045481 | 4.006298255 | 3.87<br>(3.73 to 4.01) |
| 2 | East Asia | DALYs | 3.627809843 | 3.477775603 | 3.778061621 | 3.63<br>(3.48 to 3.78) |
| 3 | Oceania | Deaths | 0.427824895 | 0.264187719 | 0.591729137 | 0.43<br>(0.26 to 0.59) |
| 4 | Oceania | DALYs | 0.316322775 | 0.143707584 | 0.489235498 | 0.32<br>(0.14 to 0.49) |
| 5 | Southeast Asia | Deaths | 2.877798968 | 2.634623027 | 3.121551075 | 2.88<br>(2.63 to 3.12) |
| 6 | Southeast Asia | DALYs | 2.581039967 | 2.314416269 | 2.848358466 | 2.58<br>(2.31 to 2.85) |
| 7 | Central Asia | Deaths | 0.295699112 | 0.17809814 | 0.413438138 | 0.3<br>(0.18 to 0.41) |
| 8 | Central Asia | DALYs | 0.059563153 | -0.056985362 | 0.176247581 | 0.06<br>(-0.06 to 0.18) |
| 9 | Central Europe | Deaths | 0.48282058 | 0.327399223 | 0.638482706 | 0.48<br>(0.33 to 0.64) |
| 10 | Central Europe | DALYs | 0.372646597 | 0.221388032 | 0.524133447 | 0.37<br>(0.22 to 0.52) |
| 11 | High-income<br>Asia Pacific | Deaths | -0.385474879 | -0.815842715 | 0.046760358 | -0.39<br>(-0.82 to 0.05) |
| 12 | High-income<br>Asia Pacific | DALYs | -0.890113498 | -1.293490131 | -0.485088415 | -0.89<br>(-1.29 to -0.49) |
| 13 | Western Europe | Deaths | 2.139643917 | 1.99419067 | 2.285304595 | 2.14<br>(1.99 to 2.29) |
| 14 | Western Europe | DALYs | 2.047396974 | 1.896219698 | 2.198798543 | 2.05<br>(1.9 to 2.2) |
| 15 | Eastern Europe | Deaths | 2.276831537 | 1.962426198 | 2.592206357 | 2.28<br>(1.96 to 2.59) |
| 16 | Eastern Europe | DALYs | 2.151033791 | 1.803468718 | 2.499785479 | 2.15<br>(1.8 to 2.5) |
| 17 | Australasia | Deaths | 4.806475594 | 4.640751268 | 4.972462386 | 4.81<br>(4.64 to 4.97) |
| 18 | Australasia | DALYs | 4.643981244 | 4.503435336 | 4.784716171 | 4.64<br>(4.5 to 4.78) |
| 19 | Southern Latin<br>America | Deaths | 4.462004773 | 4.212172052 | 4.712436431 | 4.46<br>(4.21 to 4.71) |
| 20 | Southern Latin<br>America | DALYs | 4.300291159 | 4.04440782 | 4.556803808 | 4.3<br>(4.04 to 4.56) |
| 21 | High-income<br>North America | Deaths | 3.917109407 | 3.664880328 | 4.169952191 | 3.92<br>(3.66 to 4.17) |
| 22 | High-income<br>North America | DALYs | 3.795000097 | 3.487982174 | 4.102928851 | 3.8<br>(3.49 to 4.1) |
| 23 | Caribbean | Deaths | 1.032933157 | 0.763834049 | 1.302750919 | 1.03<br>(0.76 to 1.3) |
| 24 | Caribbean | DALYs | 0.97932658 | 0.714185567 | 1.245165605 | 0.98<br>(0.71 to 1.25) |
| 25 | Andean Latin<br>America | Deaths | 1.717428965 | 1.472470975 | 1.962978291 | 1.72<br>(1.47 to 1.96) |
| 26 | Andean Latin<br>America | DALYs | 1.359922901 | 1.110813782 | 1.609645757 | 1.36<br>(1.11 to 1.61) |
| 27 | Central Latin<br>America | Deaths | 1.522098923 | 1.240204216 | 1.804778542 | 1.52<br>(1.24 to 1.8) |
| 28 | Central Latin<br>America | DALYs | 1.375518016 | 1.083864198 | 1.668013333 | 1.38<br>(1.08 to 1.67) |
| 29 | South Asia | Deaths | 5.324549643 | 5.18371838 | 5.465569466 | 5.32<br>(5.18 to 5.47) |
| 30 | South Asia | DALYs | 5.277620328 | 5.11796383 | 5.437519318 | 5.28<br>(5.12 to 5.44) |
| 31 | Tropical Latin<br>America | Deaths | 2.726776448 | 2.475273695 | 2.978896457 | 2.73<br>(2.48 to 2.98) |
| 32 | Tropical Latin America | DALYs | 2.538396394 | 2.283239096 | 2.79419021 | 2.54<br>(2.28 to 2.79) |
| 33 | Central Sub-Saharan Africa | Deaths | 2.132492874 | 2.008398505 | 2.256738206 | 2.13<br>(2.01 to 2.26) |
| 34 | Central Sub-Saharan Africa | DALYs | 2.026024961 | 1.906932469 | 2.145256629 | 2.03<br>(1.91 to 2.15) |
| 35 | North Africa and Middle East | Deaths | 2.05573718 | 1.983477753 | 2.128047806 | 2.06<br>(1.98 to 2.13) |
| 36 | North Africa and Middle East | DALYs | 2.000881343 | 1.928616853 | 2.073197066 | 2<br>(1.93 to 2.07) |
| 37 | Southern Sub-Saharan Africa | Deaths | 2.461963078 | 1.853908909 | 3.073647249 | 2.46<br>(1.85 to 3.07) |
| 38 | Southern Sub-Saharan Africa | DALYs | 2.26282309 | 1.633643807 | 2.895897408 | 2.26<br>(1.63 to 2.9) |
| 39 | Eastern Sub-Saharan Africa | Deaths | 2.388023591 | 2.261619067 | 2.514584361 | 2.39<br>(2.26 to 2.51) |
| 40 | Eastern Sub-Saharan Africa | DALYs | 2.322521406 | 2.183033701 | 2.462199523 | 2.32<br>(2.18 to 2.46) |
| 41 | Western Sub-Saharan Africa | Deaths | 1.053429792 | 0.94752632 | 1.159444367 | 1.05<br>(0.95 to 1.16) |
| 42 | Western Sub-Saharan Africa | DALYs | 0.893928712 | 0.776520324 | 1.011473885 | 0.89<br>(0.78 to 1.01) |

Abbreviations: EAPC,estimated annual percentage change; LCI,low confidence interval; UCI,up confidence interval;DALYs,disability-adjusted life-years.

The disease burden of high BMI-associated HCC was highly heterogeneous across regions. To identify regions with similar disease burdens, a hierarchical cluster analysis was conducted. The results revealed that regions such as East Asia, South Asia, high-income North America, Australia, and South America (Latin America) had significantly higher ASDR and ASR of DALYs compared to other regions. In contrast, Western Europe, Southeast Asia, and parts of sub-Saharan Africa demonstrated intermediate disease burdens, while Central Europe and high-income Asia-Pacific, regions had relatively low disease burdens (Fig 2).

**Fig 2.**
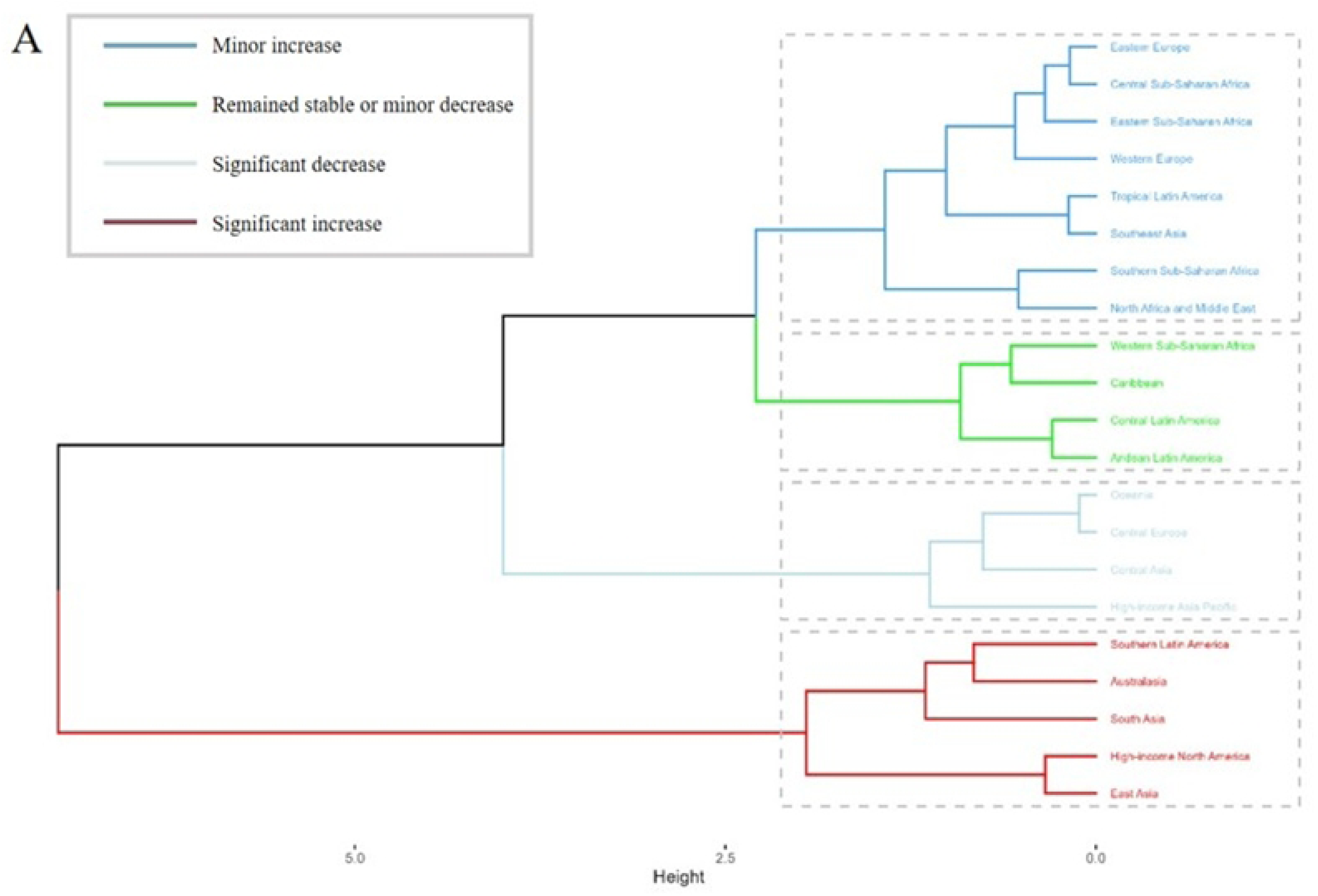
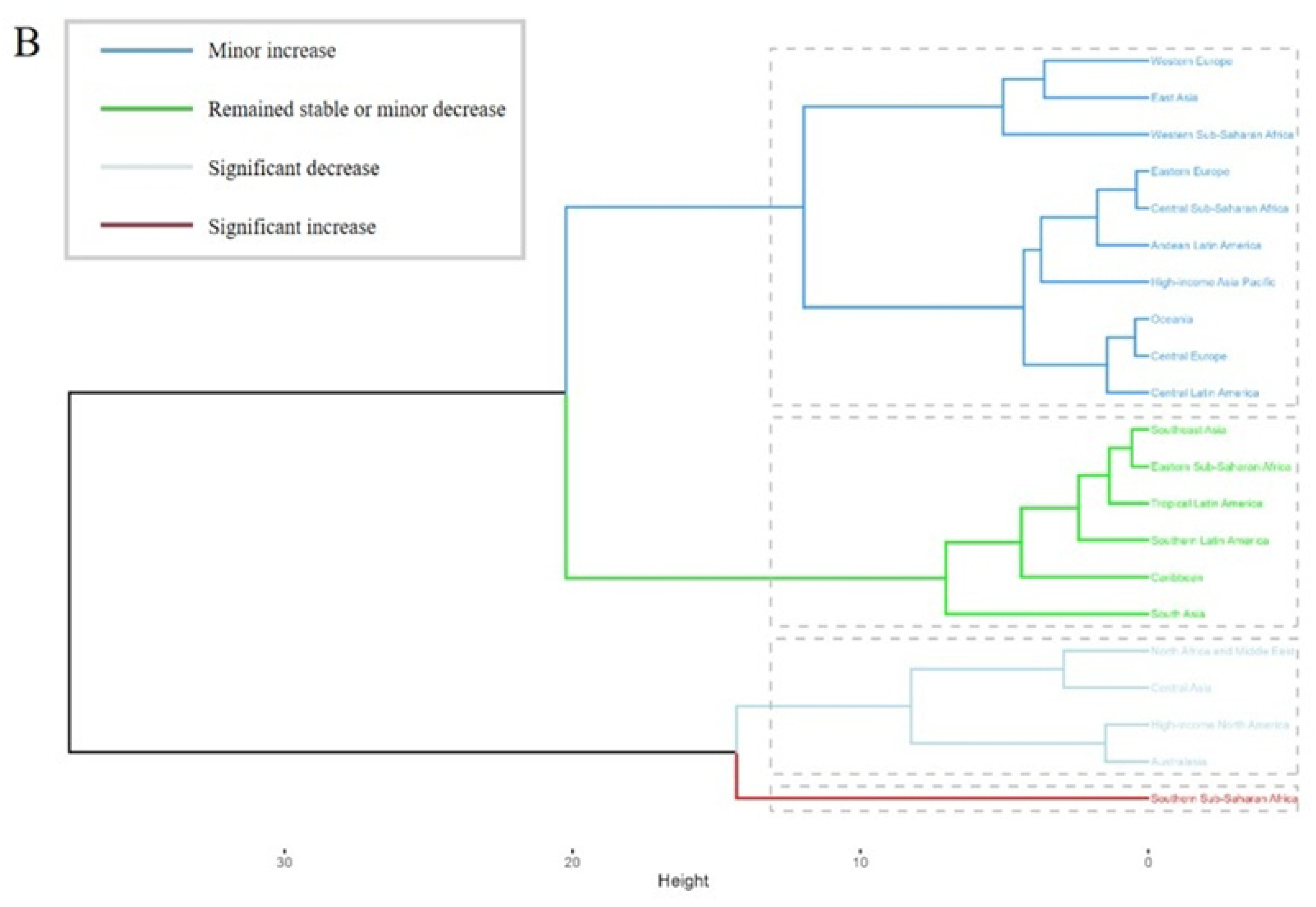
EAPC in the ASDR (A) and ASR of DALYs (B) of high BMI-associated HCC from 1990 to 2021: cluster Analysis. Abbreviations: ASDR,Age-standardized death rate;ASR,age-standardized rate;DALYs, disability-adjusted life-years.

### 3.3 Trends of high BMI-associated hepatocellular carcinoma by nations

At the national level, the burden of high BMI–associated hepatocellular carcinoma increased markedly in most urban areas. The steepest increases were observed in South Asia, where India experienced an annualized rise in mortality of 5.52% (95% CI: 5.42-5.61) and in DALYs of 5.38% (95% CI: 5.26-5.50). In Southeast Asia, DALYs grew rapidly in Indonesia (4.72%, 95% CI: 4.51-4.92) and Vietnam (4.50%, 95% CI: 4.13-4.87). East Asia presented divergent trends: China saw increases in mortality (3.84%, 95% CI: 3.68-3.99) and DALYs (3.62%, 95% CI: 3.45-3.79), whereas Japan (mortality: –0.95%, 95% CI: -1.53 to -0.37) and South Korea (DALYs: -0.70%, 95% CI: -0.86 to -0.55) experienced significant declines. Among high-income English-speaking nations, the United Kingdom (DALYs: -5.23%, 95% CI: -5.48 to -4.98) and Australia (-5.13%, 95% CI: -5.30 to -4.96) also showed decreasing DALY rates, while the United States recorded a substantial annual increase in DALYs of 3.91% (95% CI: 3.64-4.18). In summary, South and Southeast Asia and some high-income English-speaking countries (e.g., the United Kingdom, Australia) had the fastest-growing burdens, whereas some high-income East Asian nations (e.g., Japan, South Korea) showed negative growth (S1 Table).

### 3.4 Trends of high BMI-associated hepatocellular carcinoma to SDI

An analysis across SDI quintiles revealed distinct gradients in the global burden of high BMI-associated hepatocellular carcinoma from 1990 to 2021. Middle-SDI regions exhibited the most dramatic relative growth, with the ASDR climbing at an annualized rate of 3.03% (95% CI:2.94-3.13) and ASR of DALYs rising at 2.91% (95% CI:2.81-3.00), This suggests that these regions are undergoing rapid epidemiological transitions, characterized by accumulating metabolic risk factors (Table 2). Although high-SDI regions experienced more moderate annual growth (ASDR 2.52%), the high baseline burden (1990 ASDR: 0.3/100,000) led to an absolute increase of 0.4/100,000 (from 0.3 to 0.7/100,000), reflecting the long-term cumulative effect of chronic diseases.

In contrast, low-SDI regions showed the smallest growth, with the ASDR rising from 0.15/100,000 to 0.25/100,000, marking the lowest growth rate of 1.35% and an increase of +0.1/100,000. Interestingly, despite starting with the same initial ASDR as the high-middle SDI region (0.3/100,000), reached a lower final burden (0.6/100,000) owing to its higher growth rate of 3.03% (Fig 3). This spatial heterogeneity reveals that middle SDI regions are particularly influenced by emerging dietary patterns and lifestyle changes, while high SDI regions are more affected by the long-term consequences of obesity-related complications.

**Table 2.** Burden of high BMI-associated HCC in both sexes for ASR by SDI, 1990-2021.

|  | Location | measure | EAPC | LCI | UCI | EAPC(95%CI) |
| --- | --- | --- | --- | --- | --- | --- |
| 1 | High-middle SDI | Deaths | 1.989738089 | 1.903412318 | 2.076136988 | 1.99<br>(1.9 to 2.08) |
| 2 | High-middle SDI | DALYs | 2.002361771 | 1.911363017 | 2.09344178 | 2<br>(1.91 to 2.09) |
| 3 | Low-middle SDI | Deaths | 2.571341901 | 2.472711311 | 2.670067424 | 2.57<br>(2.47 to 2.67) |
| 4 | Low-middle SDI | DALYs | 2.650493314 | 2.54090282 | 2.760200933 | 2.65<br>(2.54 to 2.76) |
| 5 | High SDI | Deaths | 2.521189672 | 2.330874079 | 2.711859215 | 2.52<br>(2.33 to 2.71) |
| 6 | High SDI | DALYs | 2.338101012 | 2.133345389 | 2.543267126 | 2.34<br>(2.13 to 2.54) |
| 7 | Low SDI | Deaths | 1.346800236 | 1.267404204 | 1.426258516 | 1.35<br>(1.27 to 1.43) |
| 8 | Low SDI | DALYs | 1.358774373 | 1.279795918 | 1.437814417 | 1.36<br>(1.28 to 1.44) |
| 9 | Middle SDI | Deaths | 3.031637815 | 2.935527614 | 3.127837754 | 3.03<br>(2.94 to 3.13) |
| 10 | Middle SDI | DALYs | 2.905545222 | 2.806756262 | 3.00442911 | 2.91<br>(2.81 to 3) |

Abbreviations: EAPC,estimated annual percentage change; LCI,low confidence interval; UCI,up confidence interval;DALYs,disability-adjusted life-years.

**Fig 3.**
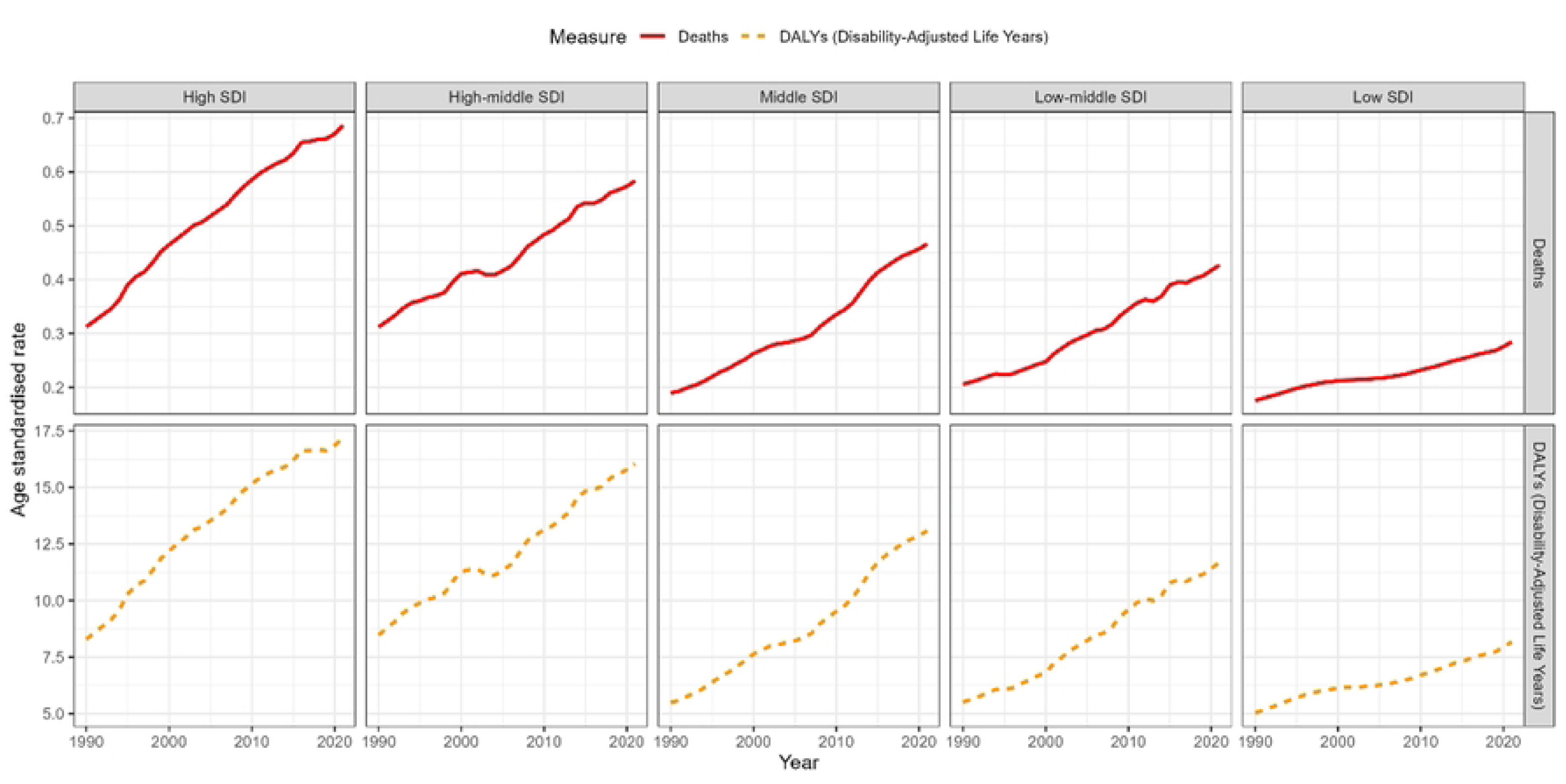
Trends in the burden of high BMI-associated HCC by SDI, 1990-2021. Abbreviations:SDI,sociodemographic index.

In addition, significant heterogeneity characterized the changes in the burden of high BMI-associated HCC across different SDI subgroups (as shown in Fig 4). In high SDI regions, such as Australasia, high-income North America, and Western Europe, both the ASDR and ASR of DALYs are relatively low and tend to decrease with increasing SDI. Middle SDI regions, such as Eastern Europe, Central Europe, and East Asia, exhibit peak disease burdens, with Eastern Europe having the highest ASDR in this group. Conversely, Low SDI regions including tropical Latin America and central and southern sub-Saharan Africa, demonstrate a striking increase in disease burden as SDI rises, despite their initially low absolute burden of disease.

**Fig 4.**
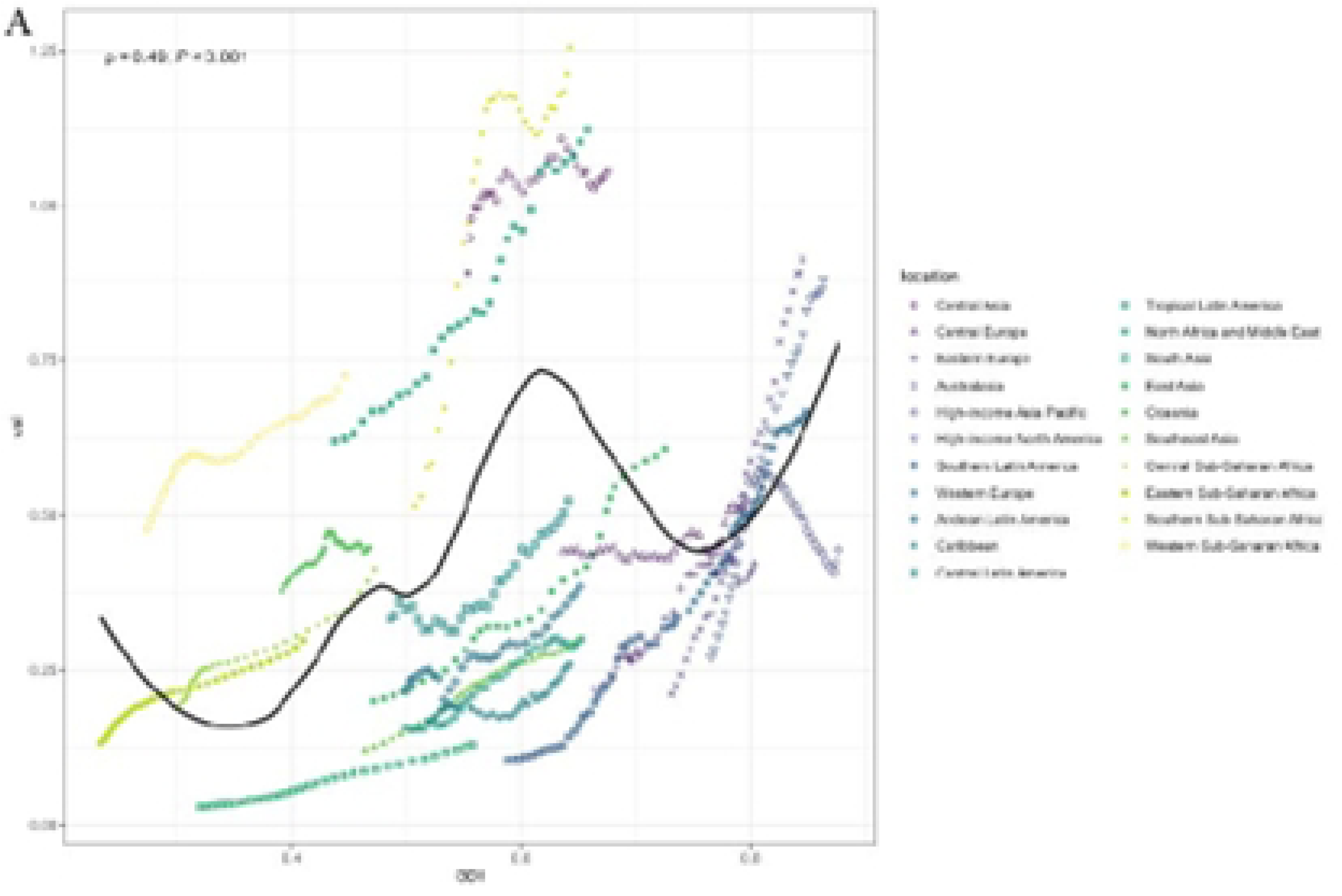
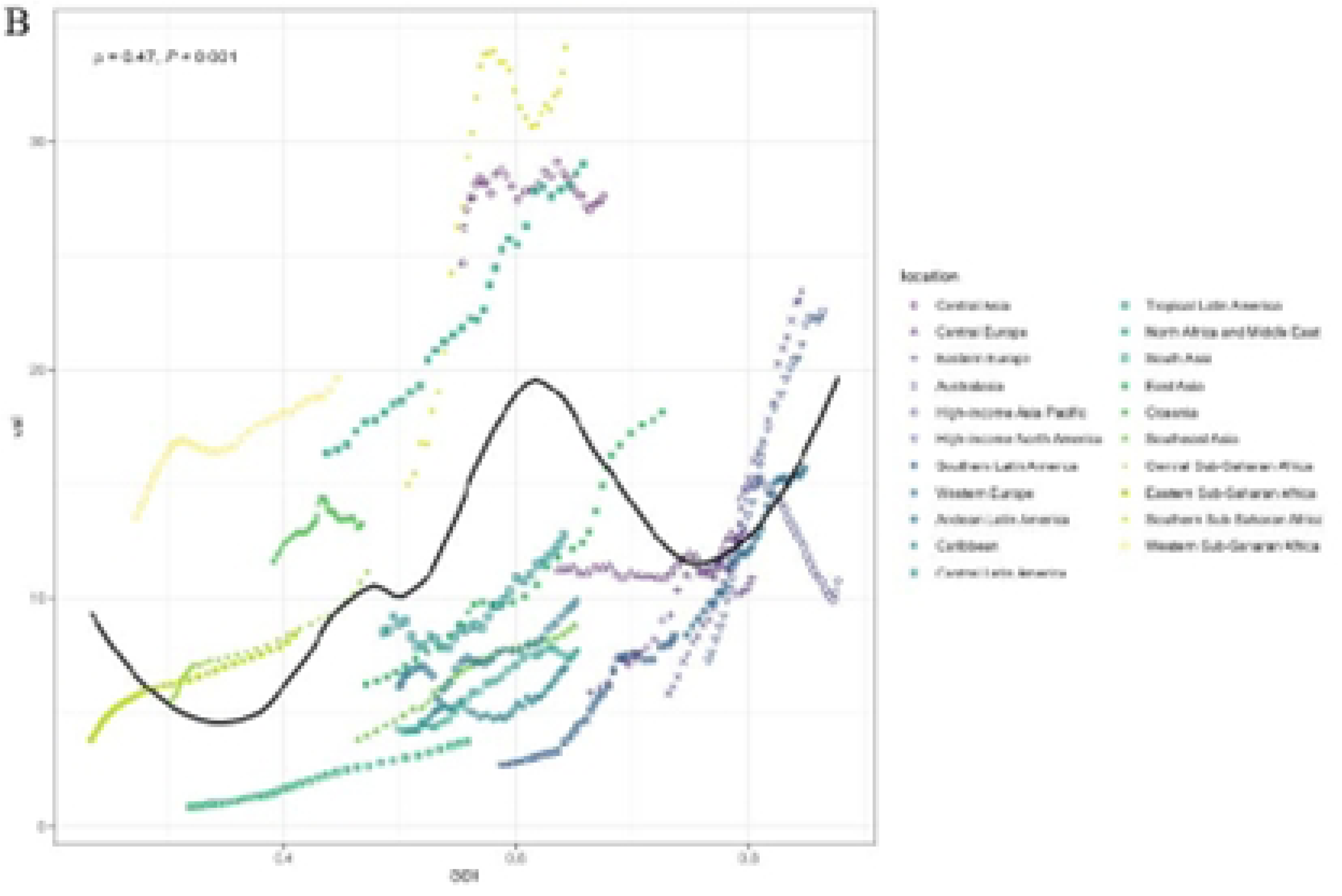
The ASDR (A) and ASR of DALYs (B) of high BMI-associated HCC across 21 GBD regions by SDI. Abbreviations: ASDR,Age-standardized death rate;ASR,age-standardized rate;DALYs, disability-adjusted life-years.SDI,sociodemographic index.

### 3.5 Gender distribution of high BMI-associated hepatocellular carcinoma

According to the GBD2021 database, the global mortality rate of high BMI-associated hepatocellular carcinoma demonstrated a significant upward trend, with an overall average annual increase of approximately 2.37% (95% CI: 2.29-2.45). Specifically, The ASDR increased at an annual rate of 2.60% (95% CI: 2.49-2.71) for males and 2.05% (95% CI: 2.00-2.09) for females, indicating a significantly higher rate of increase in males compared to females. In terms of ASR of DALYs, the overall annual average growth rate was 2.31% (95% CI: 2.22-2.39), with a growth rate of 2.51% (95% CI: 2.39-2.63) for males, again Considerably higher than the 1.97% (95% CI: 1.93-2.01) observed for females. These trends suggest that the burden of disease is rising at a faster rate in males, and females remain at risk of continued increases (Table 3).

Between 1990 and 2021, the global ASDR for high BMI–associated HCC increased markedly from 0.25 to 0.55 per 100,000. In males, the ASDR climbed from 0.30 to 0.70 per 100,000 (a 133% increase), whereas in females it increased from 0.20 to 0.40 per 100,000 (a 100% increase). Over the same period, ASR of DALYs increased from 6 to 14 per 100,000 globally. When stratified by sex, male ASR of DALYs rose from 8 to 20 per 100,000, compared with an increase from 5 to 10 per 100,000 in females. Overall, males bear a substantially higher disease burden than females, and both mortality and DALY increases were more pronounced in the male population (Fig 5).

**Table 3.** Sex differences in the burden of high BMI-associated HCC, 1990-2021.

|  | location | measure | sex | cause | age | EAPC | LCI | UCI | EAPC(95%CI) |
| --- | --- | --- | --- | --- | --- | --- | --- | --- | --- |
| 1 | Global | Deaths | Male | Liver cancer | Age-standardized | 2.596177193 | 2.486236317 | 2.706236007 | 2.6<br>(2.49 to 2.71) |
| 2 | Global | Deaths | Female | Liver cancer | Age-standardized | 2.048432452 | 2.004572268 | 2.092311496 | 2.05<br>(2 to 2.09) |
| 3 | Global | Deaths | Both | Liver cancer | Age-standardized | 2.369868322 | 2.289787133 | 2.450012206 | 2.37<br>(2.29 to 2.45) |
| 4 | Global | <sup>4</sup>DALYs | Male | Liver cancer | Age-standardized | 2.510675672 | 2.392862831 | 2.628624068 | 2.51<br>(2.39 to 2.63) |
| 5 | Global | DALYs | Female | Liver cancer | Age-standardized | 1.973116479 | 1.933077704 | 2.013170981 | 1.97<br>(1.93 to 2.01) |
| 6 | Global | DALYs | Both | Liver cancer | Age-standardized | 2.307497828 | 2.221629243 | 2.393438545 | 2.31<br>(2.22 to 2.39) |

Abbreviations: EAPC,estimated annual percentage change; LCI,low confidence interval; UCI,up confidence interval;DALYs,disability-adjusted life-years.

**Fig 5.**
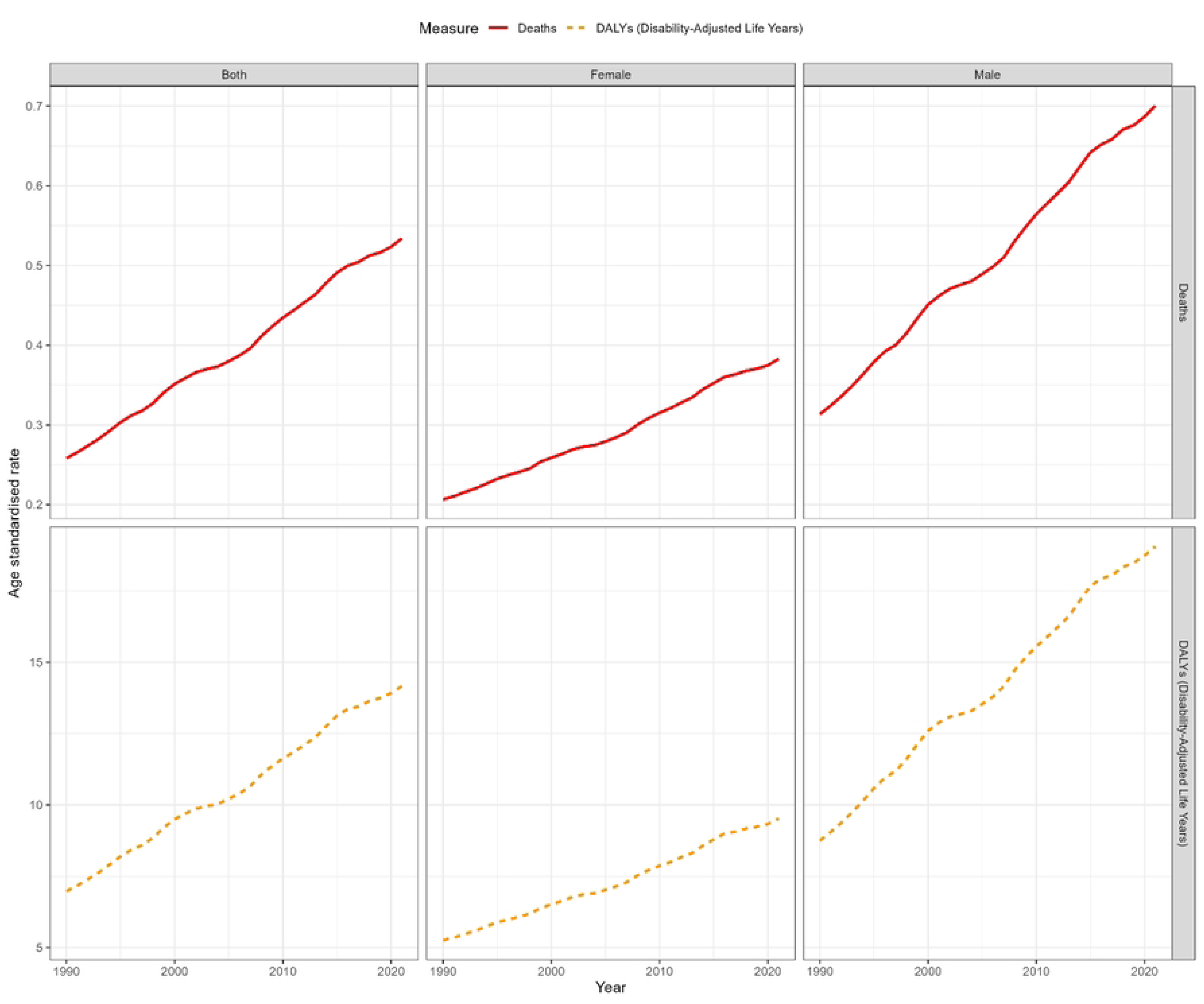
Trends in the burden of high BMI-associated HCC by gender, 1990-2021

### 3.6 Age distribution of high BMI-associated hepatocellular carcinoma

From 1990 to 2021, high BMI-associated HCC mortality and DALYs showed a continuous upward trend in all age groups, with markedly different rates of rise by age. The most pronounced increases occurred in the oldest cohorts: in those aged 95+, the average annual mortality increase was 3.12% (95% CI: 3.01–3.24) and the DALY increase was 3.07% (95% CI: 2.94–3.19); in the 80–84 age group, these rates were 2.93% and 2.92%, respectively. In contrast, younger adults (20–50 years) exhibited more moderate growth—for example, the 25–29 age bracket saw a 2.47% annual increase (95% CI: 2.19–2.75). Middle-aged to elderly adults (50–80 years) showed relatively stable growth, such as a 2.36% increase (95% CI: 2.25–2.47) in the 60–64 cohort. Overall, disease burden rose significantly with age, peaking in those ≥65 years. Specifically, in the 95+ group, the ASDR reached 96 per 100,000 and the ASR of DALYs reached 800 per 100,000. The 40–64 age group bore the second-highest burden, while those aged 20–39 years had a comparatively light burden—for instance, the ASDR in the 20–24 cohort was only 1 per 100,000 (Table 4; Fig 6).

**Table 4.**
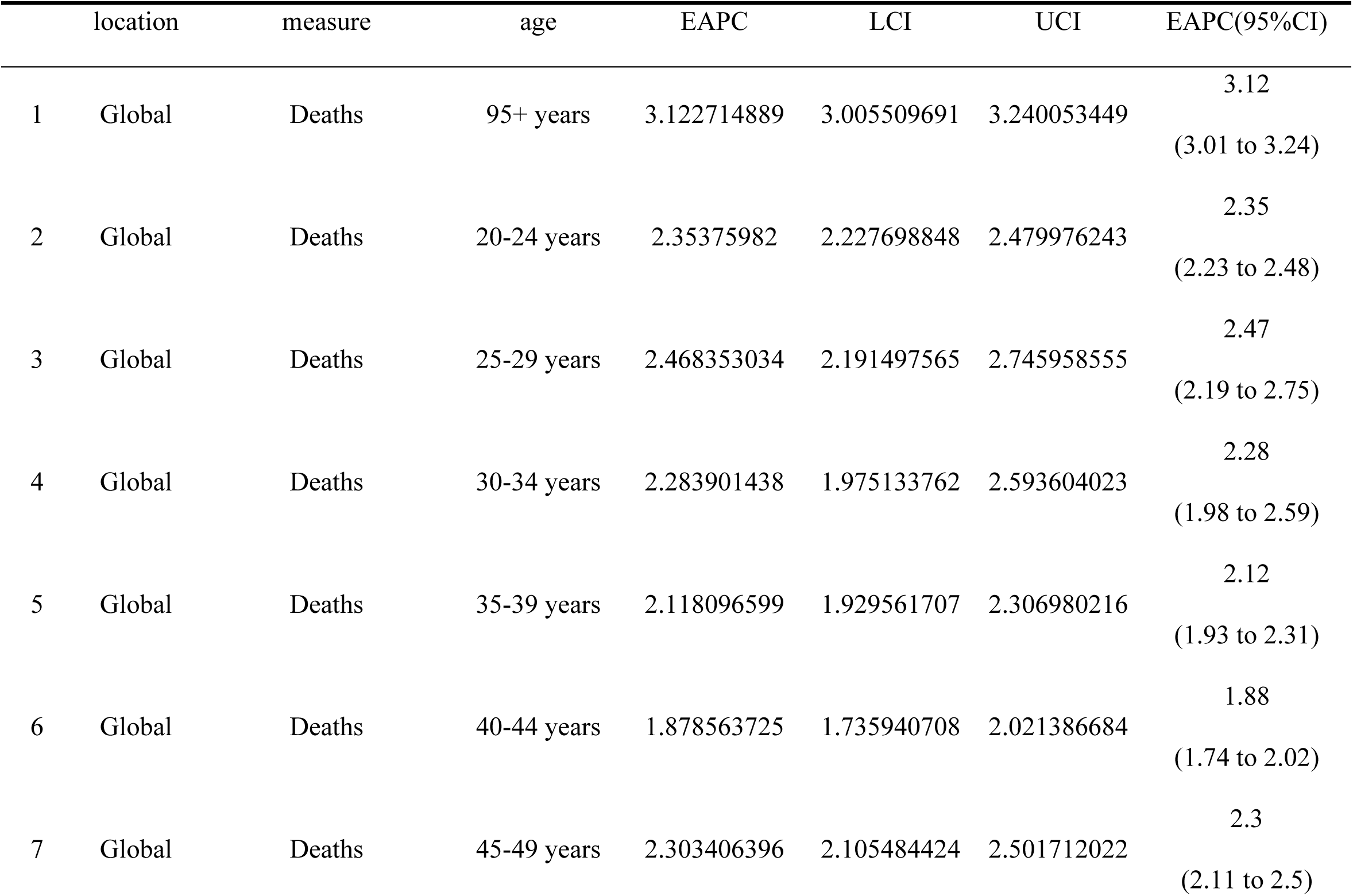
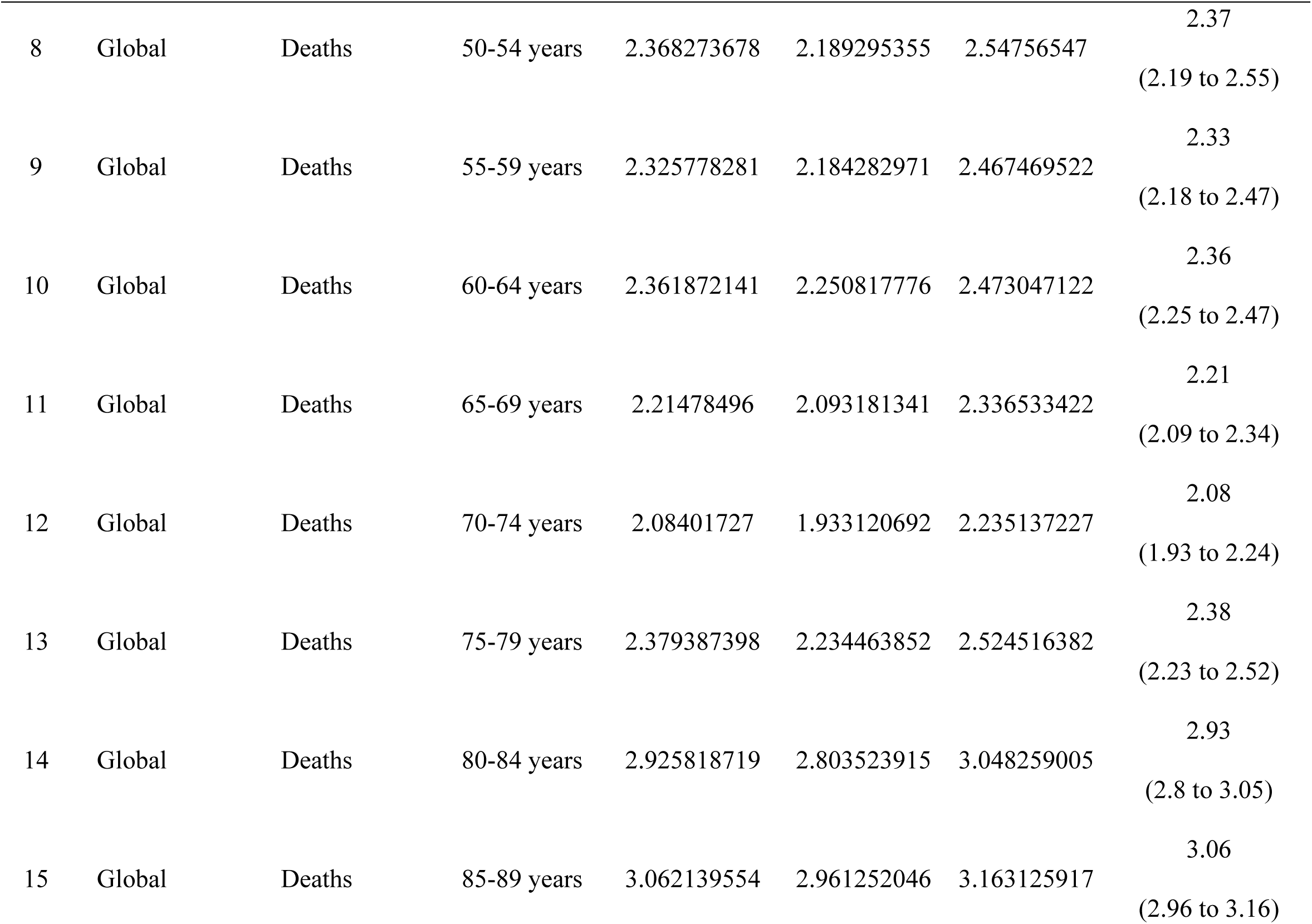
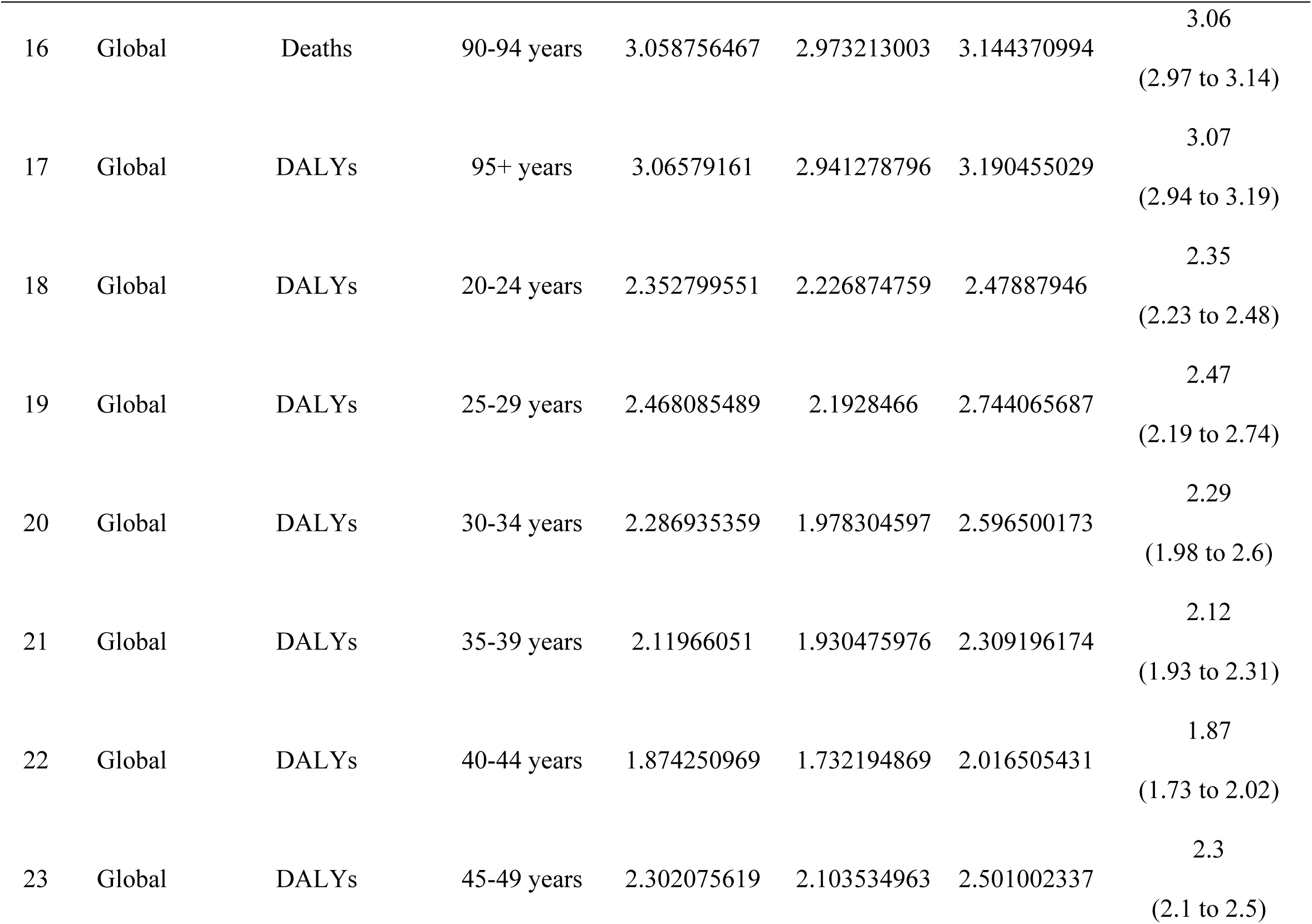
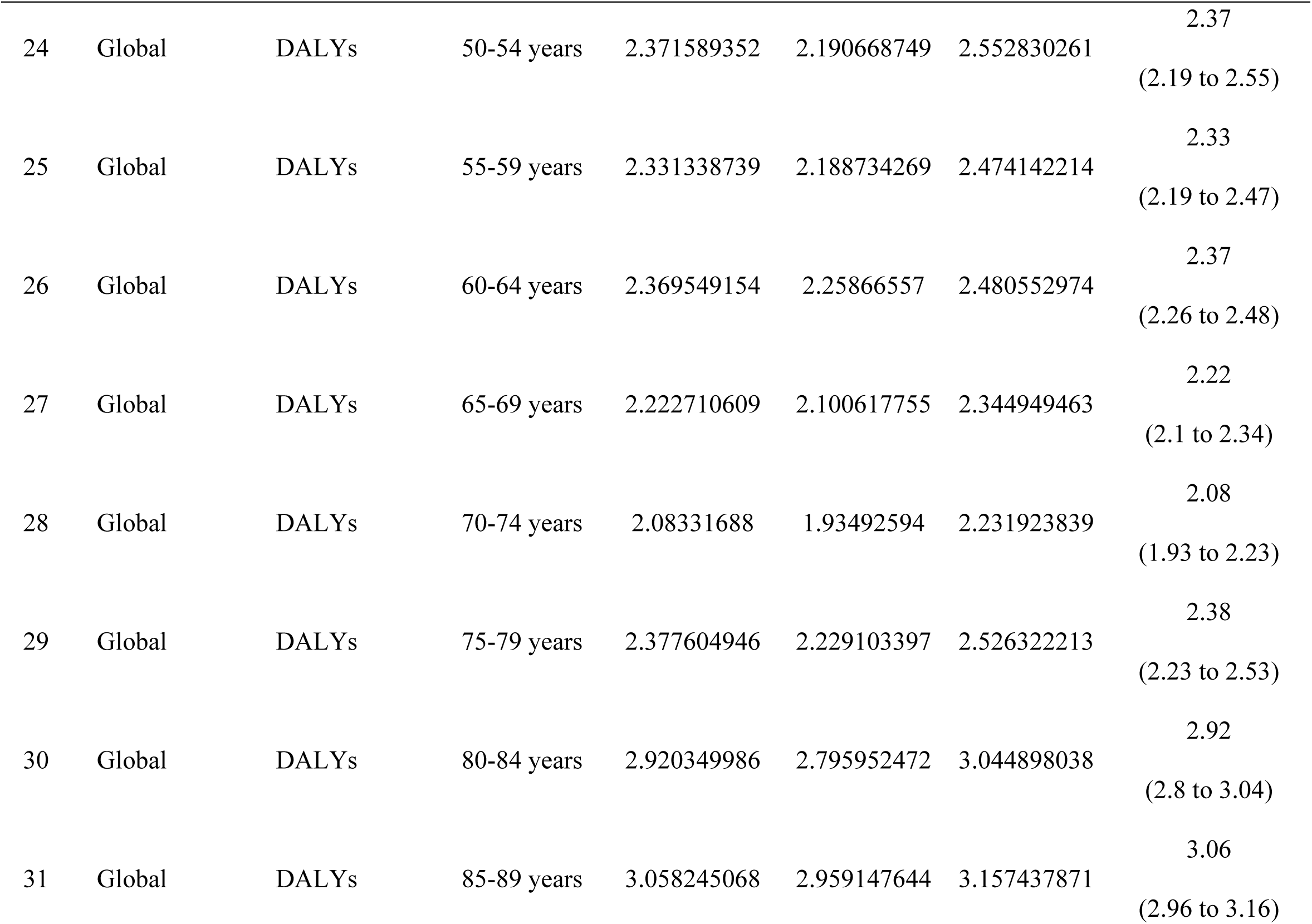
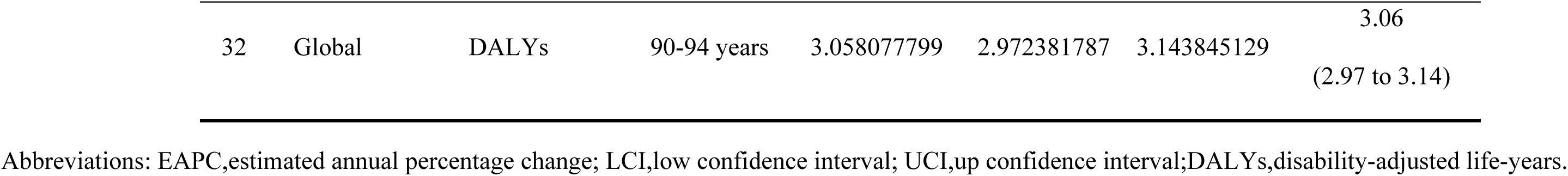
Age distribution of high BMI-associated HCC burden in both sexes, 1990-2021.

**Fig 6.**
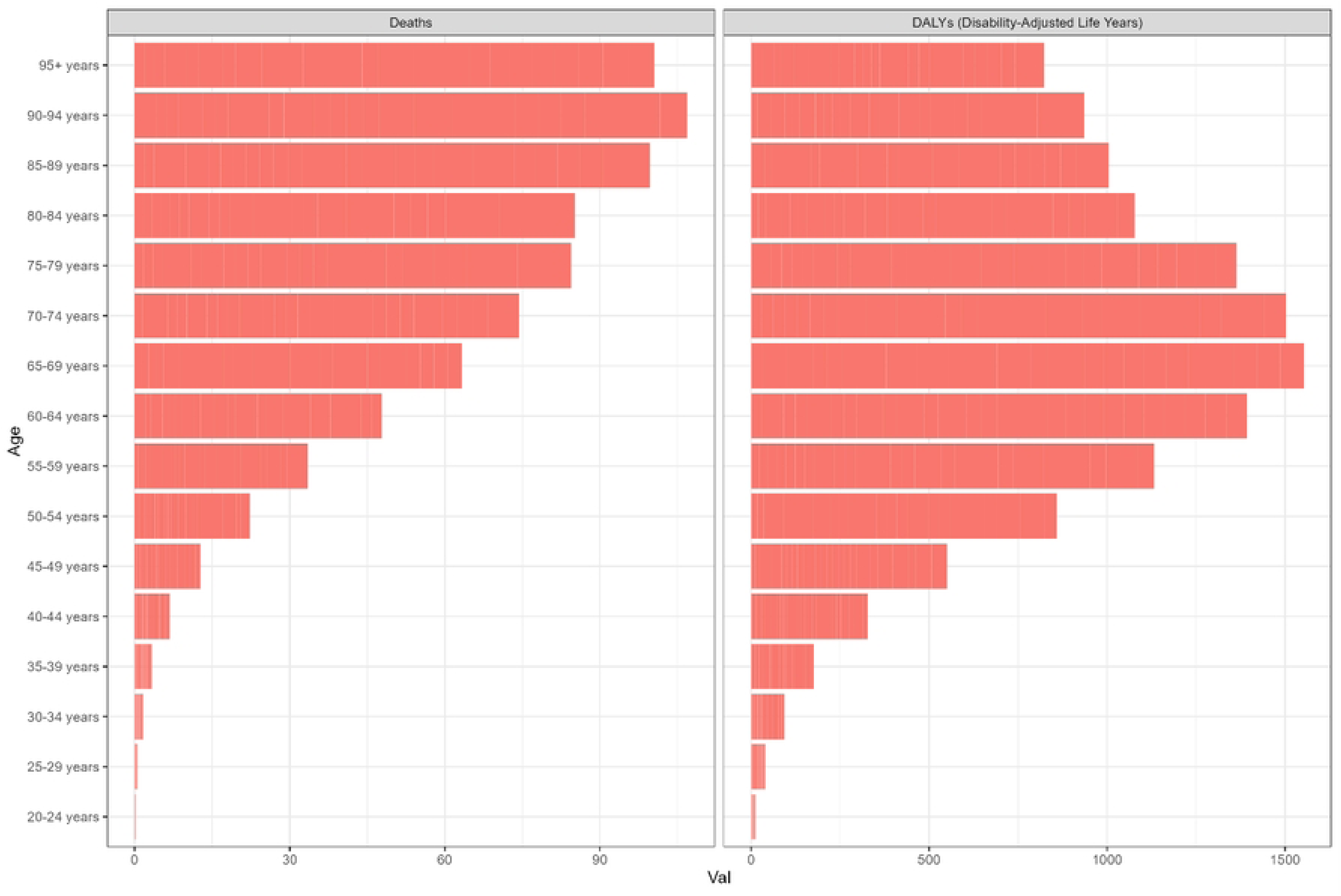
Characteristics of age distribution of high BMI-associated HCC burden, 1990-2021

### 3.7 Forecasting the burden of high BMI–associated HCC to 2040

Between 1990 and 2021, global mortality and DALYs attributable to high BMI– associated HCC increased markedly, with a more pronounced increase observed in males. Projections from 2022 to 2040 indicate that this burden will continue to grow, albeit at progressively decelerating rates. Specifically, annual mortality is projected to increase by 1.20% (95% CI: 1.15-1.25) in males and by 1.15% (95% CI: 1.10–1.20) in females, while the combined annual growth rate for DALYs is estimated at 1.13% (95% CI: 1.08-1.18) (S2 Table).

Projections using the Nordpred model revealed that ASDR for high BMI– associated HCC increased from 0.20 per 100,000 in 1990 to 0.85 per 100,000 in 2021, while age-standardized DALYs (ASR of DALYs) increased from 5 to 225 per 100,000 during the same period, These trends underscore a sustained rise in disease burden. By 2040, both ASDR and ASR of DALYs are expected to plateau, stabilizing at approximately 0.90 per 100,000 and 25 per 100,000, respectively (Fig 7).

**Fig 7.**
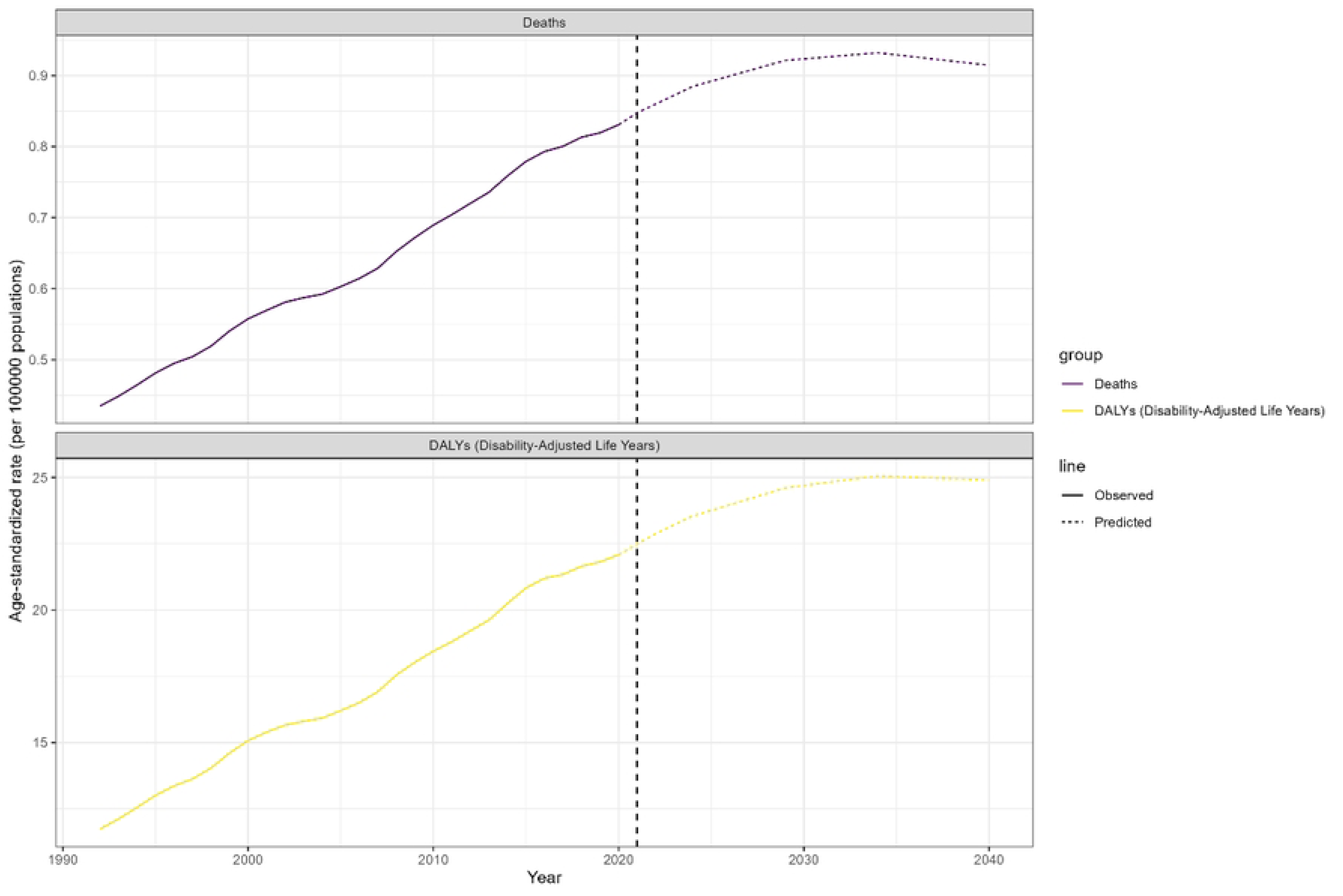
Nordpred model-based projections of high BMI-associated HCC burden, 2022-2040

## 4. Discussion

Drawing on the Global Burden of Disease Study 2021 (GBD2021), this study comprehensively quantified the global burden of high BMI-associated HCC from 1990 to 2040, and delineated temporal trends and disparities in ASDR and ASR of DALYs different across sexes, age groups, and regions. The analyses revealed that both ASDR and ASR of DALYs for high BMI-associated HCC increased substantially worldwide between 1990 and 2021., and the burden was significantly higher in males than in females. This male predominance likely reflects greater exposure to established HCC risk factors—namely, tobacco use, alcohol consumption, and hepatitis B infection— paralleling the observations of Siegel et al. [23]. Accordingly, we recommend the design and implementation of targeted, evidence-based prevention and management strategies for males to mitigate the increasing incidence of high BMI–associated HCC in this population[24].

From an age-specific perspective, the global burden of high BMI-associated HCC increases markedly with advancing age. Mortality peaks in the 90–94-year-old cohort, likely reflecting both higher background mortality in the very elderly and increased susceptibility to HCC[25], Concurrently, the prevalence of comorbid conditions such as diabetes mellitus and dyslipidemia is elevated in the elderly population, further exacerbating their overall disease burden[26]. In addition, ASR of DALYs reaches the peak in the 65-69-year-old cohort, which may be attributable to the cumulative impact of chronic health impairments that substantially compromise both quality of life and longevity. It is noteworthy that ASR of DALYs declines among those aged 70 and above, suggesting that HCC patients surviving into advanced age may experience less aggressive disease progression or achieve more effective disease control, thereby accruing fewer disability years[27–28]. These findings highlight the importance of focusing prevention and management efforts for high BMI-associated HCC on middle-aged and older adult populations.

The burden of high BMI-associated HCC exhibits significant heterogeneity across regions and nations. closely mirroring variations in the SDI as defined by the Global Burden of Disease framework. Specifically, high-SDI regions consistently experience a more severe high BMI-associated HCC burden, whereas low-to-middle SDI regions demonstrate comparatively lower rates. This trend can be attributed to the higher prevalence of overweight/obesity, diabetes mellitus, and hyperlipidemia in high-income countries, alongside the prevalence of unhealthy behaviors, such as sedentary lifestyles, excessive alcohol consumption, and high calorie diets[29]. Furthermore, countries with higher SDI levels typically face more pronounced population aging, which exacerbates their disease burden[30]. Among the different GBD super-regions, East Asia, South Asia, high-income North America, Australia, and Latin America South America consistently maintained the highest global disease burden levels between 1990 and 2021. This epidemiological pattern may be closely linked to the dietary cultures and lifestyles of these regions, including higher rates of obesity, fast-food consumption, and the prevalence of high-sugar, high-fat diets coupled with insufficient physical activity. In contrast, Central Europe and the high-income Asia Pacific region experienced relatively lower disease burdens, which can likely be attributed to healthier dietary patterns, such as diets rich in fish and vegetables and well-controlled caloric intake, as well as lower obesity rates and access to advanced medical technologies and effective health interventions[30]. Of particular note, China’s high BMI-associated HCC burden remains elevated globally, primarily due to its massive population, the accelerating process of population aging[31–32], and widespread tobacco and alcohol consumption, which collectively pose significant challenges to liver cancer prevention and control[33]. However, in recent years, China has made progress in mitigating the increasing burden of high BMI–associated HCC through the promotion of neonatal hepatitis B vaccination, adult hepatitis B screening, standardized hepatitis C treatment, and the vigorous implementation of the “Healthy China” policy[34–35]. This strongly suggests that geographical, socioeconomic, and cultural factors play a crucial role in shaping the disease burden. Given these regional disparities, flexible healthcare policies should be developed to address the unique disease burden scenarios in different regions and countries. High-SDI countries should focus on more precise early diagnosis standards, treatment protocol, and prevention strategies, while also preparing for the challenges posed by population aging. By contrast, lower-SDI countries require more international medical support to overcome challenges related to population growth and insufficient healthcare resources.

Despite the fact that this study provides valuable multidimensional insights into the global burden of high BMI–associated HCC, several limitations should be acknowledged. Primarily, our analyses rely on GBD 2021 data, the accuracy of which depends on the completeness and consistency of cancer registries and vital-registration systems worldwide, potentially introducing reporting biases. Second, the uniform diagnostic criteria for determining high BMI may not adequately consider the variations in physiological characteristics across different ethnic and regional populations. Finally, our assessment was confined to national and regional levels and did not examine subnational variations (e.g., provincial, municipal, or rural settings). Future research should aim to overcome these limitations and further enhance the understanding of the global burden of high BMI-associated HCC through more precise data collection, refined assessment criteria, and more detailed geographic segmentation. Such advancements would provide a solid scientific foundation for developing more accurate and effective public health strategies.

## 5. Conclusion

The present study confirms that high BMI-associated HCC represents a significant global burden between 1990 and 2021, with particularly pronounced impacts in middle-SDI regions. The burden is more severe in the older age group (≥ 80 years) and among males. Projections based on the Nordpred model indicate that, while the global burden of high BMI–associated HCC will continue to rise between 2022 and 2040, the growth rate is expected to decelerate gradually, with both ASDR and ASR of DALYs stabilizing by 2040. This trend suggests that high BMI-associated HCC will remain a critical challenge for global public health. When designing health policies, nations must thoroughly consider disparities in burden across different age, gender, and geographic populations. And customize flexible and effective strategies for the prevention and treatment of liver cancer for high-risk populations and high-burden areas. Healthy dietary patterns and active lifestyles should be advocated in all aspects, and early screening and targeted interventions should be strengthened,.This is essential to reduce the burden of high BMI–associated HCC. These efforts will enhance the global prevention and control of liver cancer and safeguard public health.

## Data Availability

The data underlying the results presented in the study are available from the Global Burden of Disease (GBD) 2021 database(https://ghdx.healthdata.org/gbd-results-tool).

## Acknowledgments

We express our gratitude toward the GBD 2021 collaborators and all who contributed to this study.

## Supporting information

S1 Table. Distribution of high BMI-associated hepatocellular carcinoma burden across 204 countries and regions from 1990 to 2021. (DOCX)

S2 Table. Projected global burden of high BMI-associated hepatocellular carcinoma from 2022 to 2040. (DOCX)

## References

1. Wu Z, Xia F, Lin R. Global burden of cancer and associated risk factors in 204 countries and territories, 1980-2021: a systematic analysis for the GBD 2021. J Hematol Oncol. 2024;17(1):119. Epub 20241129. doi: 10.1186/s13045-024-01640-8. PubMed PMID: 39614359; PubMed Central PMCID: PMC11607901.

2. Brown ZJ, Tsilimigras DI, Ruff SM, Mohseni A, Kamel IR, Cloyd JM, et al. Management of Hepatocellular Carcinoma: A Review. JAMA Surg. 2023;158(4):410–20. doi: 10.1001/jamasurg.2022.7989. PubMed PMID: 36790767.

3. Zheng J, Wang S, Xia L, Sun Z, Chan KM, Bernards R, et al. Hepatocellular carcinoma: signaling pathways and therapeutic advances. Signal Transduct Target Ther. 2025;10(1):35. Epub 20250207. doi: 10.1038/s41392-024-02075-w. PubMed PMID: 39915447; PubMed Central PMCID: PMC11802921.

4. Vogel A, Meyer T, Sapisochin G, Salem R, Saborowski A. Hepatocellular carcinoma. Lancet. 2022;400 (10360):1345–62. Epub 20220906. doi: 10.1016/s0140-6736(22)01200-4. PubMed PMID: 36084663.

5. 5. Collaborators GAB. Global, regional, and national prevalence of child and adolescent overweight and obesity, 1990-2021, with forecasts to 2050: a forecasting study for the Global Burden of Disease Study 2021. Lancet. 2025;405(10481):785–812. Epub 20250303. doi: 10.1016/s0140-6736(25)00397-6. PubMed PMID: 40049185; PubMed Central PMCID: PMC11920006.

6. 6. Collaborators GRF. Global burden and strength of evidence for 88 risk factors in 204 countries and 811 subnational locations, 1990-2021: a systematic analysis for the Global Burden of Disease Study 2021. Lancet. 2024;403(10440):2162–203. doi: 10.1016/s0140-6736(24)00933-4. PubMed PMID: 38762324; PubMed Central PMCID: PMC11120204.

7. 7. Collaborators GAB. Global, regional, and national prevalence of adult overweight and obesity, 1990-2021, with forecasts to 2050: a forecasting study for the Global Burden of Disease Study 2021. Lancet. 2025;405(10481):813–38. Epub 20250303. doi: 10.1016/s0140-6736(25)00355-1. PubMed PMID: 40049186; PubMed Central PMCID: PMC11920007.

8. Saha A, Kolonin MG, DiGiovanni J. Obesity and prostate cancer - microenvironmental roles of adipose tissue. Nat Rev Urol. 2023;20(10):579–96. Epub 20230517. doi: 10.1038/s41585-023-00764-9. PubMed PMID: 37198266.

9. Escala-Garcia M, Morra A, Canisius S, Chang-Claude J, Kar S, Zheng W, et al. Breast cancer risk factors and their effects on survival: a Mendelian randomisation study. BMC Med. 2020;18(1):327. Epub 20201117. doi: 10.1186/s12916-020-01797-2. PubMed PMID: 33198768; PubMed Central PMCID: PMC7670589.

10. Paragomi P, Zhang Z, Abe SK, Islam MR, Rahman MS, Saito E, et al. Body Mass Index and Risk of Colorectal Cancer Incidence and Mortality in Asia. JAMA Netw Open. 2024;7(8):e2429494. Epub 20240801. doi: 10.1001/jamanetworkopen.2024.29494. PubMed PMID: 39196559; PubMed Central PMCID: PMC11358861.

11. Recalde M, Pistillo A, Davila-Batista V, Leitzmann M, Romieu I, Viallon V, et al. Longitudinal body mass index and cancer risk: a cohort study of 2.6 million Catalan adults. Nat Commun. 2023;14(1):3816. Epub 20230630. doi: 10.1038/s41467-023-39282-y. PubMed PMID: 37391446; PubMed Central PMCID: PMC10313757.

12. Wu C, Targher G, Byrne CD, Mao Y, Cheung TT, Yilmaz Y, et al. Global, Regional, and National Burden of Primary Liver Cancer Attributable to Metabolic Risks: An Analysis of the Global Burden of Disease Study 1990-2021. Am J Gastroenterol. 2025. Epub 20250103. doi: 10.14309/ajg.0000000000003288. PubMed PMID: 39749919.

13. Bray F, Laversanne M, Sung H, Ferlay J, Siegel RL, Soerjomataram I, et al. Global cancer statistics 2022: GLOBOCAN estimates of incidence and mortality worldwide for 36 cancers in 185 countries. CA Cancer J Clin. 2024;74(3):229-63. Epub 20240404. doi: 10.3322/caac.21834. PubMed PMID: 38572751.

14. Collaborators GCoD. Global burden of 288 causes of death and life expectancy decomposition in 204 countries and territories and 811 subnational locations, 1990-2021: a systematic analysis for the Global Burden of Disease Study 2021. Lancet. 2024;403(10440):2100–32. Epub 20240403. doi: 10.1016/s0140-6736(24)00367-2. PubMed PMID: 38582094; PubMed Central PMCID: PMC11126520.

15. Collaborators GDaI. Global burden of 369 diseases and injuries in 204 countries and territories, 1990-2019: a systematic analysis for the Global Burden of Disease Study 2019. Lancet. 2020;396(10258):1204–22. doi: 10.1016/s0140-6736(20)30925-9. PubMed PMID: 33069326; PubMed Central PMCID: PMC7567026.

16. Hu W, Yang J. Effect of ambient ozone pollution on disease burden globally: A systematic analysis for the global burden of disease study 2019. Sci Total Environ. 2024;926:171739. Epub 20240319. doi: 10.1016/j.scitotenv.2024.171739. PubMed PMID: 38508259.

17. Cao F, Xu Z, Li XX, Fu ZY, Han RY, Zhang JL, et al. Trends and cross-country inequalities in the global burden of osteoarthritis, 1990-2019: A population-based study. Ageing Res Rev. 2024;99:102382. Epub 20240623. doi: 10.1016/j.arr.2024.102382. PubMed PMID: 38917934.

18. Global, regional, and national disability-adjusted life-years (DALYs) for 315 diseases and injuries and healthy life expectancy (HALE), 1990-2015: a systematic analysis for the Global Burden of Disease Study 2015. Lancet. 2016;388(10053):1603–58. doi: 10.1016/s0140-6736(16)31460-x. PubMed PMID: 27733283; PubMed Central PMCID: PMC5388857.

19. Ding Q, Liu S, Yao Y, Liu H, Cai T, Han L. Global, Regional, and National Burden of Ischemic Stroke, 1990-2019. Neurology. 2022;98(3):e279–e90. Epub 20211215. doi: 10.1212/wnl.0000000000013115. PubMed PMID: 34911748.

20. Xie Z, Yu C, Cui Q, Zhao X, Zhuang J, Chen S, et al. Global Burden of the Key Components of Cardiovascular-Kidney-Metabolic Syndrome. J Am Soc Nephrol. 2025. Epub 20250305. doi: 10.1681/asn.0000000658. PubMed PMID: 40042920.

21. Wang K, Chen S, Wang M, Han Q, Hou Y, Wang X. Global, regional, and National Burden of chronic kidney disease attributable to dietary risks from 1990 to 2021. Front Nutr. 2025;12:1555159. Epub 20250325. doi: 10.3389/fnut.2025.1555159. PubMed PMID: 40201583; PubMed Central PMCID: PMC11975581.

22. Bray F, Møller B. Predicting the future burden of cancer. Nat Rev Cancer. 2006;6(1):63–74. doi: 10.1038/nrc1781. PubMed PMID: 16372017.

23. Siegel RL, Miller KD, Wagle NS, Jemal A. Cancer statistics, 2023. CA Cancer J Clin. 2023;73(1):17–48. doi: 10.3322/caac.21763. PubMed PMID: 36633525.

24. Xing QQ, Li JM, Chen ZJ, Lin XY, You YY, Hong MZ, et al. Global burden of common cancers attributable to metabolic risks from 1990 to 2019. Med. 2023;4(3):168–81.e3. Epub 20230302. doi: 10.1016/j.medj.2023.02.002. PubMed PMID: 36868237.

25. Liu C, Zhu S, Zhang J, Wu P, Wang X, Du S, et al. Global, regional, and national burden of liver cancer due to non-alcoholic steatohepatitis, 1990-2019: a decomposition and age-period-cohort analysis. J Gastroenterol. 2023;58(12):1222–36. Epub 20230904. doi: 10.1007/s00535-023-02040-4. PubMed PMID: 37665532.

26. Aunan JR, Watson MM, Hagland HR, Søreide K. Molecular and biological hallmarks of ageing. Br J Surg. 2016;103(2):e29–46. doi: 10.1002/bjs.10053. PubMed PMID: 26771470.

27. Forner A, Reig M, Bruix J. Hepatocellular carcinoma. Lancet. 2018;391 (10127): 1301–14. Epub 20180105. doi: 10.1016/s0140-6736(18)30010-2. PubMed PMID: 29307467.

28. Yang X, Yang C, Zhang S, Geng H, Zhu AX, Bernards R, et al. Precision treatment in advanced hepatocellular carcinoma. Cancer Cell. 2024;42(2):180–97. doi: 10.1016/j.ccell.2024.01.007. PubMed PMID: 38350421.

29. Xing QQ, Li JM, Dong X, Zeng DY, Chen ZJ, Lin XY, et al. Socioeconomics and attributable etiology of primary liver cancer, 1990-2019. World J Gastroenterol. 2022;28(21):2361–82. doi: 10.3748/wjg.v28.i21.2361. PubMed PMID: 35800181; PubMed Central PMCID: PMC9185214.

30. Liu X, Yang W, Petrick JL, Liao LM, Wang W, He N, et al. Higher intake of whole grains and dietary fiber are associated with lower risk of liver cancer and chronic liver disease mortality. Nat Commun. 2021;12(1):6388. Epub 20211104. doi: 10.1038/s41467-021-26448-9. PubMed PMID: 34737258; PubMed Central PMCID: PMC8568891.

31. Fang EF, Scheibye-Knudsen M, Jahn HJ, Li J, Ling L, Guo H, et al. A research agenda for aging in China in the 21st century. Ageing Res Rev. 2015;24(Pt B):197–205. Epub 20150822. doi: 10.1016/j.arr.2015.08.003. PubMed PMID: 26304837; PubMed Central PMCID: PMC5179143.

32. Chen X, Giles J, Yao Y, Yip W, Meng Q, Berkman L, et al. The path to healthy ageing in China: a Peking University-Lancet Commission. Lancet. 2022;400(10367):1967–2006. Epub 20221121. doi: 10.1016/s0140-6736(22)01546-x. PubMed PMID: 36423650; PubMed Central PMCID: PMC9801271.

33. Li Q, Cao M, Lei L, Yang F, Li H, Yan X, et al. Burden of liver cancer: From epidemiology to prevention. Chin J Cancer Res. 2022;34(6):554–66. doi: 10.21147/j.issn.1000-9604.2022.06.02. PubMed PMID: 36714347; PubMed Central PMCID: PMC9829497.

34. Tanaka M, Katayama F, Kato H, Tanaka H, Wang J, Qiao YL, et al. Hepatitis B and C virus infection and hepatocellular carcinoma in China: a review of epidemiology and control measures. J Epidemiol. 2011;21(6):401–16. Epub 20111022. doi: 10.2188/jea.je20100190. PubMed PMID: 22041528; PubMed Central PMCID: PMC3899457.

35. Wang Y, Zhao L, Gao L, Pan A, Xue H. Health policy and public health implications of obesity in China. Lancet Diabetes Endocrinol. 2021;9(7):446–61. Epub 20210604. doi: 10.1016/s2213-8587(21)00118-2. PubMed PMID: 34097869.

